# Multimodal Transformer Modeling of Rapamycin Treatment in Alzheimer’s Disease via Random Forest Feature Filtering

**DOI:** 10.64898/2026.08.22.26361114

**Authors:** Chris Wang, Carter Woods, Thong Nguyen, Jian Liu, Ai-Ling Lin, Jianlin Cheng

## Abstract

Alzheimer’s Disease (AD) remains a leading cause of cognitive decline with no known cure, motivating the development of therapies that slow neurodegeneration. Rapamycin, an FDA-approved inhibitor of the mammalian target of rapamycin (mTOR) pathway, has demonstrated promising anti-aging and neuroprotective effects. However, characterizing its treatment effects and identifying the biological factors that contribute to treatment response remain challenging because of complex interactions across multiple biological systems and the limited availability of patient data. In this work, we propose a three-stage multimodal deep learning framework called TreatmentFormer for predicting rapamycin treatment status from heterogeneous biomedical data including both brain imaging data and tabular data (e.g., microbiome profiles, blood-based biomarkers, cerebral blood flow measurements, and clinical variables (e.g., gender, age, and body mass index)). First, a Random Forest-based feature selection module reduces noise in high-dimensional tabular data while preserving representation across modalities. Second, modality-specific encoders map imaging and tabular inputs into a shared latent space via self-supervised contrastive learning, enabling alignment across modalities. Finally, a transformer-based architecture integrates these representations to capture cross-modal interactions and perform treatment classification. Evaluated on a cohort of 23 participants with baseline and post-treatment timepoints, TreatmentFormer achieves an average prediction accuracy of 71.25% across 10 independent test runs. Despite the challenges of small sample size and heterogeneous data, the model demonstrates stable and consistent performance. Post hoc SHAP-based feature analysis further identifies key biomarkers associated with treatment response, particularly within blood-based and inflammatory modalities. These findings demonstrate that combining feature selection with multimodal representation learning provides a promising and robust approach for modeling treatment effects in small-sample biomedical studies. Importantly, this framework may have significant implications for clinical research and medical applications by identifying the biological features and quantitative measurements that drive individual responses to rapamycin. Such insights could facilitate the development of predictive biomarkers, improve patient stratification, and ultimately inform future approaches to AD diagnosis and therapeutic development.

## 1. Introduction

Alzheimer’s Disease (AD) is the most common form of dementia and a leading cause of death among individuals aged 65 and older, affecting approximately 50 million people worldwide [1]. Despite extensive research, effective interventions that target early disease mechanisms remain limited, highlighting the need for therapies that address underlying biological processes rather than solely managing symptoms [1].

Rapamycin (sirolimus), an FDA-approved inhibitor of mammalian target of rapamycin (mTOR) pathway, has emerged as a promising candidate due to its anti-aging and neuroprotective properties [2]. mTOR plays a central role in regulating cellular growth, metabolism, and autophagy, and its inhibition has been shown to extend lifespan in animal models while improving vascular function, mitochondrial activity, and protein homeostasis [3]. In human studies, low-dose mTOR inhibition has demonstrated potential cognitive and immunological benefits, including improved cognitive performance and enhanced immune responses [4]. Additionally, rapamycin has been shown to influence neurovascular function, peripheral metabolism, and microbiome composition, making it a compelling candidate for therapeutic intervention in neurodegenerative diseases [5].

Despite its promise, understanding the effects of rapamycin treatment remains challenging due to the complex and multifactorial nature of AD [6]. Treatment response is influenced by a combination of genetic, metabolic, inflammatory, and vascular factors, many of which interact in nonlinear ways [4]. This complexity motivates the use of machine learning approaches capable of integrating heterogeneous data sources [7].

Traditional computational approaches in AD research have primarily focused on diagnostic classification tasks, such as distinguishing between cognitively unimpaired individuals, mild cognitive impairment, and AD [8]. While these approaches have achieved strong performance, they do not directly address treatment response, which is inherently more subtle and difficult to detect [9]. Unlike diagnostic labels, treatment effects often manifest as small, time-dependent changes within the same individual, making them more susceptible to noise and variability [10].

Previously, we developed APOEFormer [11], a multimodal deep learning framework designed to classify the presence of the apolipoprotein E *ε*4 (APOE4) allele. The model employed modality-specific encoders to process heterogeneous data sources and was pretrained using self-supervised contrastive learning to capture meaningful cross-modal relationships. The resulting latent representations were concatenated and passed into a transformer architecture, which models interactions across modalities. The transformer output was then fed into a classification head to predict APOE4 status. APOEFormer obtained a 75% accuracy of predicting if a patient is a APOE4 carrier or not. This genotype prediction model helps elucidate some key features relevant such as blood metabolites to the presence of APOE4 allele.

In this work, we further extend the deep learning model used with APOEFormer to predict rapamycin treatment, aiming to identify key features that may be modulated by the treatment. Specifically, we define a binary prediction task: given multimodal data from a subject at a specific timepoint, predict whether the subject has received rapamycin treatment. In this work, each subject contributes two samples corresponding to baseline (prior to treatment) and post-treatment measurements, introducing additional challenges related to temporal variation and potential data leakage.

To address these challenges, we propose a multimodal deep learning framework called TreatmentFormer (Figure 1) that combines Random Forest-based feature filtering with the APOEFormer base model [11], which includes contrastive representation learning [12] and transformer-based integration [13]. Deep learning methods have emerged as strong options in biomedical data integration, and methods such as convolutional neural networks have been widely applied to MRI-based AD classification [14], while multilayer perceptrons (MLPs) and autoencoder-based architectures have been used to model tabular clinical and biomarker data [15]. The Random Forest module that does not exist in the APOEFormer base model is used in the initial feature selection to reduce noise in high-dimensional tabular data while maintaining modality diversity, prior to applying deep learning to them. A self-supervised contrastive learning [12] stage aligns modality-specific embeddings into a shared latent space, and a transformer-based model captures cross-modal interactions to perform classification.

**Figure 1.**
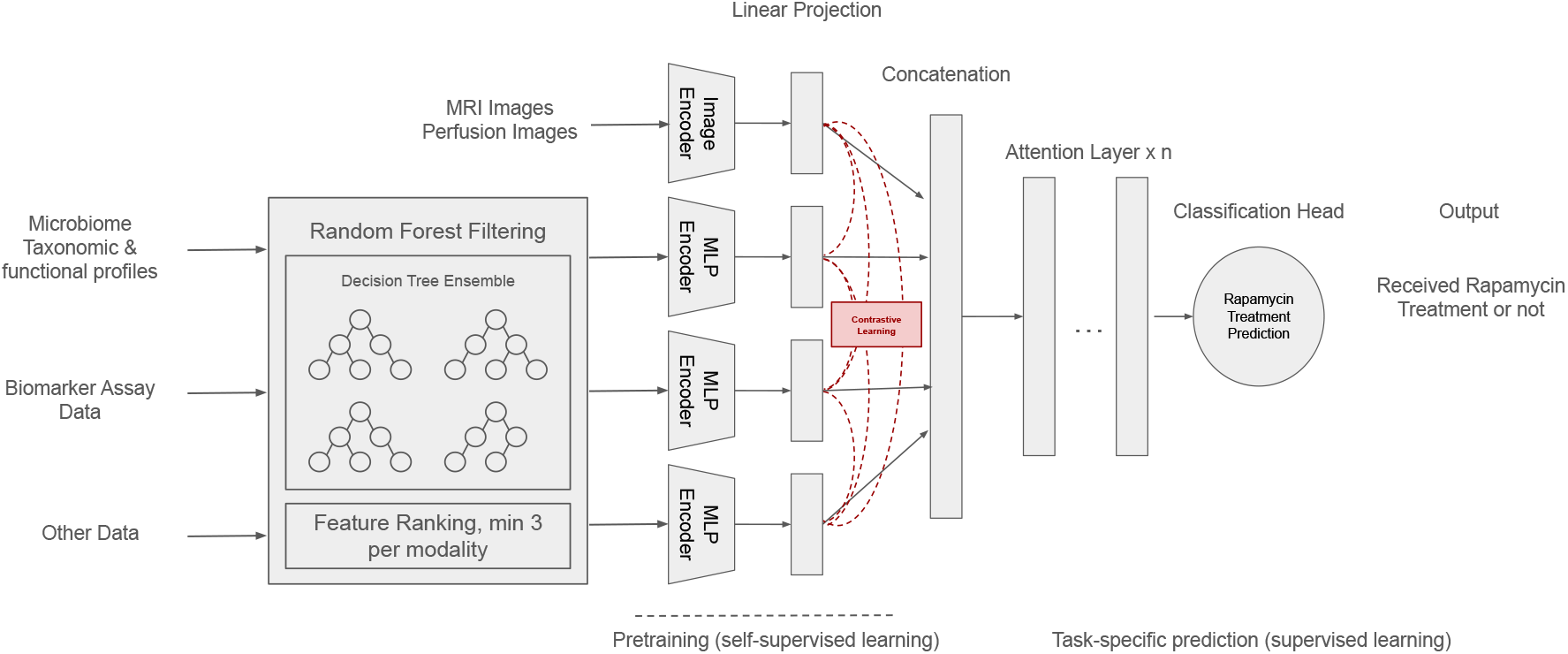
Overview of TreatmentFormer - a three-stage multimodal AI framework for Rapamycin treatment classification with random forest-based feature filtering.

This approach is designed specifically for small-sample, high-dimensional biomedical settings, where traditional deep learning methods often struggle [10]. By combining feature filtering with multimodal representation learning, the TreatmentFormer aims to improve both robustness and interpretability in modeling treatment effects. Importantly, the framework can also identify key biological features and quantitative measurements that drive individual responses to rapamycin treatment. These insights may have significant clinical implications by facilitating the identification of predictive biomarkers and informing patient stratification, AD diagnosis, and the development of more targeted therapeutic strategies.

## 2. Materials and Methods

### 2.1. Model Overview

The workflow of TreatmentFormer, a three-stage multimodal framework for modeling rapamycin treatment effects from heterogeneous biomedical data, is illustrated in Figure 1. The model builds upon the APOEFormer architecture, with key modifications to incorporate Random Forest-based feature filtering and new treatment-specific prediction.

The first stage focuses on reducing noise in the high-dimensional tabular data using Random Forest feature selection. The most informative features are retained while ensuring representation across modalities. In the second stage, modality-specific encoders transform raw inputs into latent embeddings. These embeddings are aligned in a shared latent space using a self-supervised contrastive learning objective, enabling the model to capture relationships across modalities at the subject level. In the third stage, the pretrained embeddings are treated as tokens and passed into a transformer-based model, which integrates information across modalities through self-attention mechanisms to produce a subject-level prediction of treatment status.

### 2.2. Stage 1: Random Forest-based Feature Filtering

When applying multimodal deep learning models to small biomedical datasets, a key challenge arises from the combination of high-dimensional inputs and limited sample size. In this setting, models are highly susceptible to overfitting and may struggle to distinguish meaningful patterns from noise. In our initial experiments, directly applying the APOEFormer base architecture to the treatment classification task without feature filtering resulted in near-random performance, with an average test accuracy of approximately 50%. This indicates that the model was unable to extract a reliable signal from the full set of input features due to high data dimensionality and small sample size. This failure is largely due to the presence of a large number of weak or noisy features, particularly within tabular modalities. These features increase the dimensionality of the input space, making it difficult for the transformer to effectively learn cross-modal interactions [16]. As a result, the model’s attention mechanism becomes diluted across irrelevant inputs, limiting its ability to focus on informative signals. To address this, we add a feature filtering stage using Random Forest, designed to reduce noise while preserving the most informative features across modalities, which is described below in detail.

#### 2.2.1. Random Forest-based Feature Filtering

Random Forest (RF) is an ensemble learning method composed of multiple decision trees trained using bootstrap sampling and feature subsampling [17]. At each split, a random subset of features is considered, and splits are selected to maximize the reduction in impurity, typically measured using Gini impurity. Feature importance is computed by aggregating the total impurity reduction contributed by each feature across all trees. This provides a natural and interpretable ranking of features based on their contribution to predictive performance.

RF is particularly well-suited for high-dimensional biomedical tabular data, where features are numerous, heterogeneous, and often noisy [16]. Its use of bagging reduces variance and improves stability, making it robust in small-sample settings [17]. In addition, feature importance provides valuable insight to what passes through the filter, and helps see what individual features the filter deems important to the classification task [17].

Alternative feature selection methods, such as LASSO or principal component analysis (PCA), were considered but not adopted due to their limitations in this context [18]. LASSO enforces sparsity through linear regularization, which may fail to capture nonlinear relationships between features that are common in biomedical data. PCA, while effective for dimensionality reduction, produces transformed features that are difficult to interpret biologically and may obscure modality-specific information. Gradient boosting methods such as XGBoost offer strong predictive performance but can produce unstable feature importance rankings in small datasets due to their sequential learning process and sensitivity to hyperparameters [19]. In contrast, Random Forest provides a balance of robustness, interpretability, and stability. Its feature importance estimates are less sensitive to overfitting and better reflect generalizable patterns, making it well-suited for feature filtering in small-sample multimodal settings.

### 2.2.2. Feature Selection Strategy

In TreatmentFormer, the tabular features (i.e., non-imaging features such as microbiome profiles, blood-based biomarkers, cerebral blood flow measurements, and clinical variables) are fed through Random Forest, and following computation of feature importance scores, the top 27 tabular features were selected for downstream modeling. This threshold was determined empirically, with performance observed to plateau in the range of 25–35 features.

To ensure that the multimodal nature of the dataset is preserved, an additional constraint was imposed: at least three features from each modality must be included in the selected set. This prevents the model from collapsing onto a single dominant modality and encourages learning of cross-modal interactions.

This design choice is particularly important in multimodal transformer architectures, where the ability to model relationships across modalities is central to performance [20]. By enforcing modality diversity, the model is better positioned to leverage complementary information from heterogeneous data sources.

### 2.2.3. Impact on Downstream Transformer Learning

Feature filtering provides several key benefits for the downstream transformer model.

First, reducing the number of input features minimizes the influence of irrelevant or weak signals [16]. Without filtering, the model must implicitly learn to ignore noise, which can hinder training and reduce generalization performance.

Second, high-dimensional inputs increase model variance, particularly in small datasets. This increases the risk of overfitting to spurious correlations rather than learning meaningful patterns [21]. Third, transformers compute pairwise interactions across all input tokens. As the number of tokens increases, the attention space grows quadratically, making it more difficult for the model to identify salient relationships [13]. By restricting the input to the most informative features, the model can more effectively allocate attention and learn meaningful cross-modal interactions.

Together, these effects lead to the substantial improvement in performance observed when incorporating Random Forest feature filtering into the deep learning pipeline.

### 2.3. Stage 2: Modality Encoding and Alignment

#### 2.3.1. Image feature encoding

Structural MRI and perfusion image volumes provide rich anatomical and functional information but present challenges due to their high dimensionality and variability across subjects. To address these challenges, a slice-wise encoding strategy leverages large-scale pretrained visual representations generated by a pretrained vision-language model, CLIP ViT-B/32 [22], while preserving depth-wise anatomical structure.

Each 3D volume is decomposed along the axial dimension into an ordered sequence of 2D slices. This enables the use of pretrained 2D vision models while preserving depth-wise anatomical structure. Each slice is normalized to account for low-contrast regions and missing values, then converted into a three-channel image format. Feature embeddings are generated from the 2D slices using the pretrained CLIP ViT-B/32 model, leveraging its strong transferability from large-scale image-text pretraining to medical imaging tasks [22]. Given the limited dataset size, a gradual unfreezing strategy is applied, where layers are progressively fine-tuned based on validation performance.

The resulting high-dimensional embeddings generated by the CLIP ViT-B/32 model are projected into a compact latent space using a lightweight projection head consisting of linear transformations, normalization, and nonlinear activation. The final embeddings (hidden features) are normalized to ensure consistent scale and organized as a depth-ordered sequence, with padding applied to enable batch processing.

#### 2.3.2. Tabular feature encoding

In addition to imaging data, the model incorporates multiple tabular modalities, including microbiome profiles, blood-based biomarkers, cerebral blood flow measurements, and clinical variables (e.g., gender, age, and body mass index [BMI]) [5]. As described in Stage 1, 27 features are selected from these modalities by the random forest module, with at least three features for each tabular modality.

These tabular modalities differ in dimensionality and statistical distribution, which can lead to scale imbalance and suboptimal feature interactions. To address this, each modality is given a dedicated multi-layer perceptron (MLP) encoder that maps its raw input features into a shared latent embedding space.

Each encoder consists of stacked transformation blocks composed of linear projection, layer normalization, nonlinear activation, and dropout. A final normalization step ensures that all modality embeddings lie on a comparable scale, enabling stable interactions during multimodal fusion [23].

#### 2.3.3. Self-Supervised Contrastive Representation Learning for Modality Alignment

A key challenge in multimodal biomedical data integration is aligning heterogeneous modalities that differ in scale, noise characteristics, and semantic meaning. To address this, a contrastive learning-based pretraining strategy is used to align modality-specific embeddings at the subject level (Figure 1).

The objective encourages embeddings from different modalities of the same subject to be similar, while embeddings from different subjects are pushed apart. This is implemented using an InfoNCE-style loss [24], where positive pairs consist of embeddings from the same subject across modalities, and negative pairs consist of embeddings from different subjects [24].

Pairwise similarities are computed using temperature-scaled cosine similarity. The contrastive objective promotes compact clustering of embeddings belonging to the same subject while maintaining separation across subjects. This alignment allows the model to learn consistent cross-modal representations prior to downstream classification. The encoders and projection layers were trained by the contrastive learning in a self-supervised mode because no patient labels are required in the pretraining phase.

### 2.4. Stage 3: Transformer-based Data Integration and Treatment Prediction

#### 2.4.1. Multimodal Transformer Integration

Following self-supervised contrastive pretraining in Stage 2, modality-specific embeddings are treated as tokens and passed into a transformer-based architecture for treatment classification. Transformer models are particularly well-suited for this task due to their ability to capture complex dependencies through attention mechanisms [13]. By dynamically weighting features and modeling long-range interactions, transformers can effectively integrate heterogeneous information without relying on fixed receptive fields.

Each transformer layer consists of multi-head self-attention followed by residual connections, dropout, and layer normalization. Through stacked attention layers, the model learns higher-order interactions across modalities and adaptively reweights their contributions based on relevance to the prediction task.

To obtain a fixed-dimensional representation, the variable-length token sequence is aggregated using a learned attention pooling mechanism [25]. This approach assigns an importance score to each token and computes a weighted sum of token embeddings, enabling the model to focus on the most informative modalities for each subject.

### 2.4.2. Supervised Learning for Treatment Prediction

The transformer model is fine-tuned in a supervised manner to predict treatment status. Each subject at a time point (prior to treatment or post-treatment) is represented as a sequence of modality embeddings and processed by the transformer to produce a classification output.

To prevent data leakage, training is conducted using patient-level splits, ensuring that samples from the same individual do not appear across training and evaluation sets. Optimization is performed using the AdamW optimizer with weight decay, gradient clipping, and mixed-precision training to improve stability and computational efficiency.

To address class imbalance and improve robustness, a focal loss objective is employed [26]. Focal loss extends binary cross-entropy by introducing a modulating factor that down-weights well-classified examples and places greater emphasis on harder, misclassified samples.

### 2.5. Experimental Setup

#### 2.5.1. Dataset

The multimodal biomedical dataset used in this work consists of heterogeneous data collected from 23 patients at risk of dementia in Columbia, Missouri, aged between 40 and 60 years [5]. The dataset reflects the multifactorial nature of neurodegenerative disease progression and includes both imaging and tabular modalities.

A total of eight modalities were used, including two imaging modalities (structural MRI, perfusion imaging) and six tabular modalities (inflammatory markers, blood metabolites, cerebral blood flow (CBF) measurements, microbiome data, complete blood count (CBC), and clinical/demographic variables). Structural MRI provides detailed anatomical information, while perfusion imaging (obtained using arterial spin labeling (ASL)) captures functional blood flow dynamics. Structural MRI has been widely used for quantifying neurodegeneration and regional atrophy patterns in aging and neurological disorders [27]. ASL is a non-invasive MRI technique that quantifies cerebral blood flow (CBF) by magnetically labeling inflowing arterial blood water, enabling voxel-wise perfusion mapping without contrast agents [28]. The tabular modalities capture systemic biological processes, including metabolism, inflammation, and host-microbiome interactions.

Each patient contributes two samples corresponding to pre-treatment (Baseline) and post-treatment (Post) timepoints. As a result, the dataset contains paired samples per subject, enabling analysis of treatment-induced changes over time. This is a main difference in data preprocessing compared to the APOEFormer model for APOE4 allele prediction in our previous study [11]. In the APOE4 classification task, the genotype is fixed per patient, which means the patient’s APOE classification doesn’t change overtime or as a result of the rapamycin treatment. However, in this work, the same patient has two treatment status at two different time points (prior to rapmycin treatment and post-treatment).

#### 2.5.2. Data Partitioning and Data Leakage Prevention

To evaluate the proposed framework rigorously, experiments were conducted using patient-level data splits to prevent data leakage. The dataset was partitioned into 16 patients for training, 3 for validation, and 4 for testing. Because each patient contributes both baseline and post-treatment samples, the test set contains 8 total samples per run. Patient-level partitioning was necessary because having same patient data split between the training and testing set can lead to inflated results due to same patient data memorization and the model learning patterns that are not intrinsic to the treatment impact.

To further improve robustness, 10 independent experimental runs were performed with different randomized patient splits. The union of the test sets of the 10 runs cover all 23 patients. This repeated evaluation strategy provides a more reliable estimate of model performance in small-sample settings.

Model performance was measured using classification accuracy. Due to the small size of the test set, accuracy changes in discrete increments of 12.5%, corresponding to a single misclassification. This granularity is important when interpreting variability across runs.

## 3. Results

### 3.1. Results of Treatment Prediction with/without Feature Filtering

To assess the impact of feature filtering, the TreatmentFormer architecture was first evaluated without Random Forest preprocessing. Under this setting, the model achieved an average test accuracy of approximately 50% across 10 runs, with results distributed around chance-level performance.

This outcome suggests that the model is unable to effectively learn from the full set of high-dimensional inputs. The presence of noisy or weak features likely overwhelms the model’s ability to identify meaningful patterns, reinforcing the need for feature selection in this setting.

After introducing Random Forest-based feature filtering, the model demonstrates a substantial improvement in performance. Across 10 independent runs, the model achieves a much higher average test accuracy of 71.25% (Figure 2).

**Figure 2.**
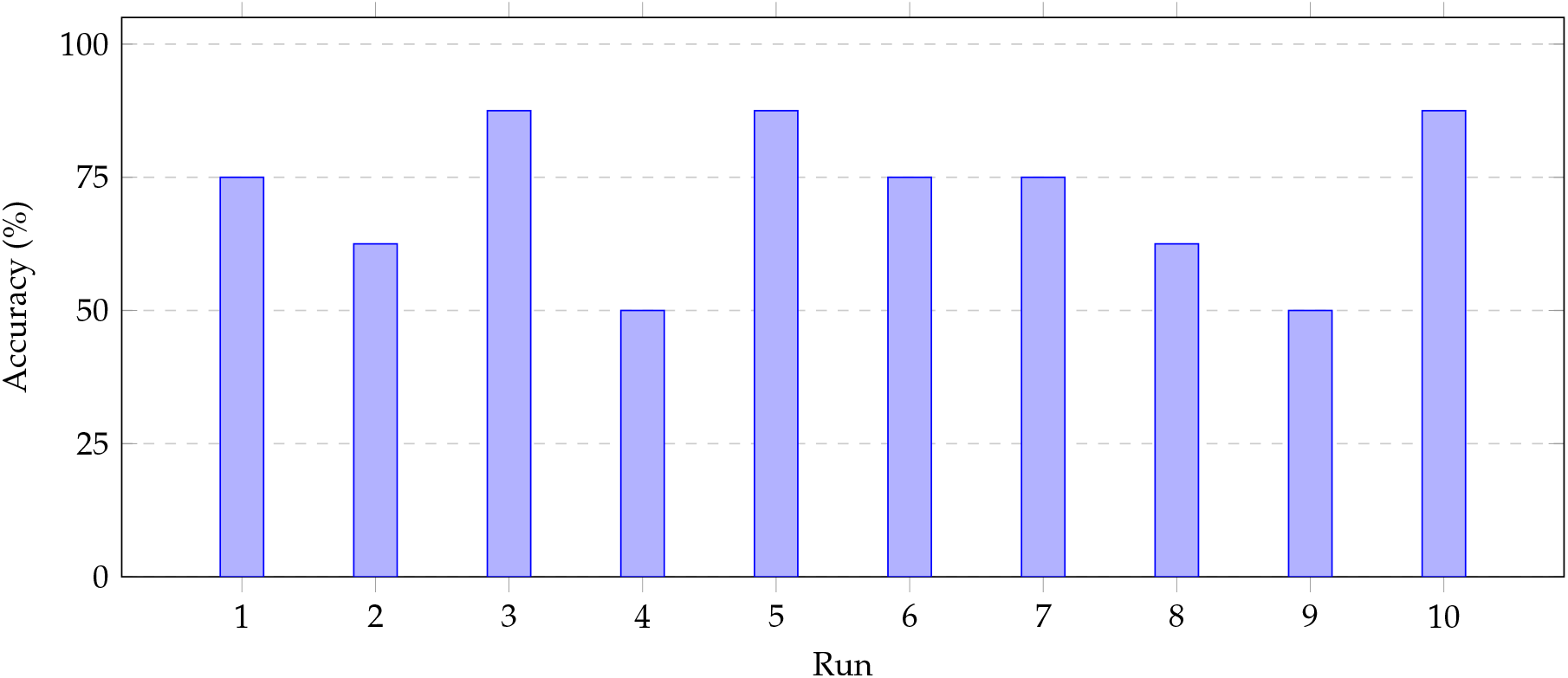
Rapamycin Treatment classification accuracy across 10 independent test runs. Performance variability reflects differences in data partitioning and sample composition across runs.

The distribution of results includes: - 3 runs at 75% accuracy - 3 runs at 87.5% accuracy - 2 runs at 62.5% accuracy - 2 runs at 50% accuracy. Given the small test size (8 samples) of each test run, these values correspond to discrete increments based on the number of correct predictions. Despite this limitation, the results remain relatively stable across runs, with accuracy extremes at 50% and 87.5%.

Importantly, the union of test sets across runs covers all 23 subjects, ensuring that evaluation is not dependent on a single fixed partition. This provides a more comprehensive assessment of model generalization. The observed improvement in performance highlights the importance of feature filtering in small-sample multimodal learning. By reducing input dimensionality and removing noisy features, the model is better able to focus on informative signals and learn meaningful cross-modal relationships.

These findings reinforce the hypothesis that controlling feature space is critical for enabling transformer-based models to perform effectively in high-dimensional, low-sample biomedical settings.

### 3.2. Random Forest-based Feature Ranking

Feature importance scores generated by the Random Forest feature filtering were aggregated across the 10 runs to identify consistently informative tabular features (see the top 27 consensus features and their modalities in Figure 3). The most dominant tabular features included RDW CV % from complete blood count and MIP-1*α* from inflammatory markers, indicating that hematological and inflammatory signals play a significant role in distinguishing treatment status.

**Figure 3.**
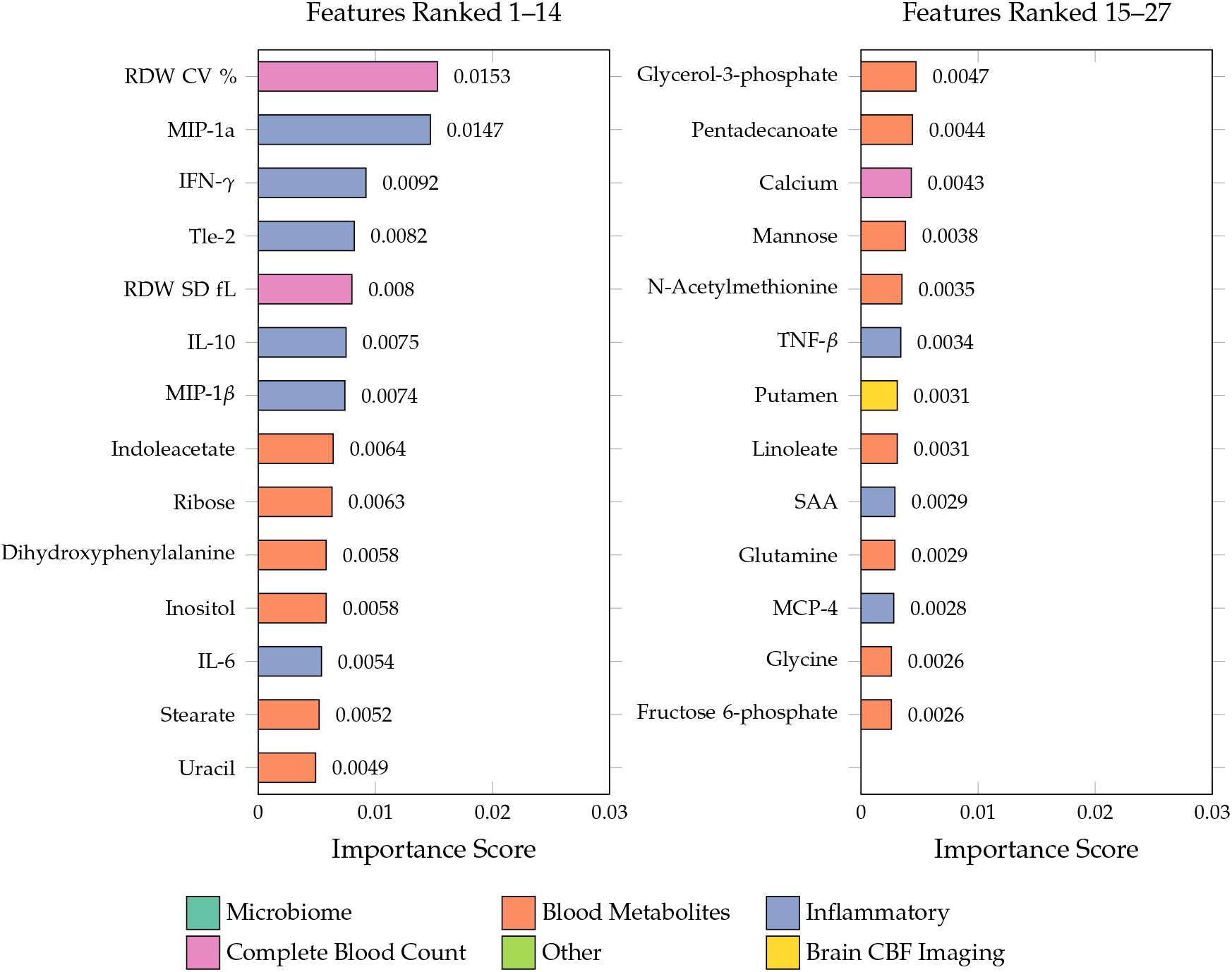
Top 27 features ranked by importance, colored by modality.

Across the top 27 tabular features, the majority originated from blood-based biomarker modalities, with particularly strong representation from inflammatory markers and metabolite profiles (Figure 3). Although CBC and Inflammatory dominated the top 7 features, blood metabolites had the most features in the top 27 with 14 features, meaning that while individual features weren’t strong, the modality as a whole plays a large role. In contrast, microbiome and demographic features were not consistently selected, suggesting that their contribution to the short-term treatment prediction may be more subtle.

Interestingly, only one feature was selected from the Brain CBF modality (Putamen), but the feature exhibited moderate importance ranked at 21. This suggests that while this modality may be less dense in informative features, certain signals within it are highly relevant.

### 3.3. SHAP-based Feature Analysis

The feature important analysis in the previous section is based on Random Forest-based feature section prior to applying the deep learning model to use the selected features to predict treatment. To quantify the contribution of each modality and each feature when all the selected tabular features and imaging features are used together with the deep learning model, SHAP (SHapley Additive exPlanations) analysis was performed on the test data during each run (Figure 4). Modality-level importance scores were computed by aggregating mean absolute SHAP values across tokens in each modality and averaging results over 10 independent runs.

**Figure 4.**
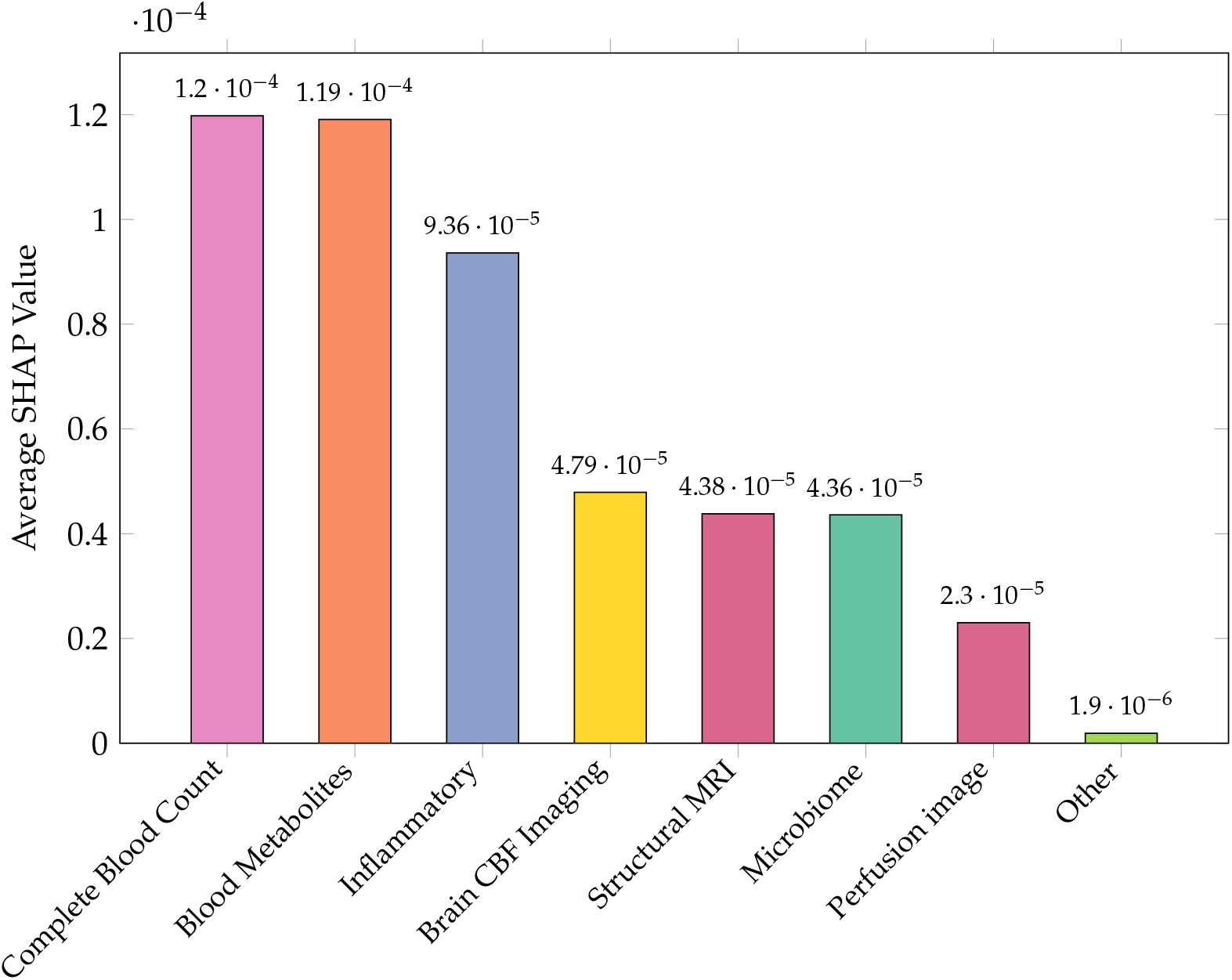
Average absolute SHAP values across 8 input modalities, indicating the relative contribution of each modality to model predictions when they are integrated together.

The results indicate that blood-based modalities, including complete blood count and metabolite profiles, consistently exhibit the highest importance. Inflammatory markers, particularly those associated with treatment response, also contribute strongly to the model’s predictions. These findings suggest that systemic biological signals play a dominant role in distinguishing baseline and post-treatment states, which is consistent with the feature analysis based on the Random Forest feature filtering.

The structural MRI and CBF value modalities demonstrates moderate importance. While the modalities may not be the primary drivers of classification, they provide complementary information that enhances predictive performance when integrated with blood-based and inflammatory tabular data.

Microbiome features also exhibit meaningful contributions, indicating a potential link between host-microbe interactions and treatment response. Perfusion imaging demonstrates less importance, suggesting that its contribution may be more subtle or context-dependent within this modeling framework.

In contrast, demographic and clinical variables show significantly low importance, which is expected given that these features are largely static and less reflective of short-term treatment effects. While Random Forest feature selection directly influences the input feature space, SHAP analysis reflects the behavior of the downstream transformer model. The consistency between Random Forest feature rankings and SHAP importance suggests that the filtering stage successfully preserves biologically meaningful signals.

Importantly, the transformer is still able to model interactions across modalities beyond the selected features, indicating that feature filtering does not limit the model’s capacity but rather improves its ability to focus on relevant information.

## 4. Discussion

This study demonstrates that combining Random Forest-based feature filtering with multimodal deep learning enables effective modeling of treatment response in small-sample biomedical datasets. The substantial improvement in performance observed after feature filtering highlights the importance of controlling input dimensionality when applying transformer-based architectures in high-dimensional, small-sample settings.

Across 10 independent test runs, the proposed framework achieves a mean classification accuracy of 71.25%, with relatively consistent performance despite variability in data partitions. This stability suggests that the model captures generalizable patterns associated with treatment effects rather than overfitting to specific subsets of patients.

A key finding is the dominant role of complete blood count (CBC) biomarkers in predicting treatment status. Both the top feature in the Random Forest feature ranking and the top modality in the SHAP analysis belonged to CBC, indicating that systemic biological processes are strongly associated with rapamycin treatment response. In addition, tabular biomarker modalities such as Blood Metabolites and Inflammatory markers had a strong presence in Random Forest and SHAP analysis. CBF tabular data, while not as dominant as other tabular biomarker counterparts, provided useful information, which was evident in one feature represented in the top 27 and the fourth highest modality SHAP value. Imaging modalities and microbiome, while less dominant, provide complementary information that enhances overall model performance. Lastly, demographic clinical data yielded the lowest SHAP values, possibly revealing the relative unimportance of the modality.

These results highlight the importance of multimodal integration, where different data sources contribute distinct but complementary signals. The ability of the transformer to capture cross-modal interactions further reinforces the value of combining heterogeneous modalities in biomedical modeling.

More broadly, these findings have potential implications for the application of multimodal artificial intelligence in medical research and precision medicine. Beyond predicting treatment status, the interpretability of TreatmentFormer provides an opportunity to identify the biological features and quantitative measurements that contribute most strongly to individual responses to rapamycin. Such information may help uncover biomarkers of treatment response and provide insight into the biological mechanisms underlying therapeutic effects. With validation in larger and more diverse cohorts, this approach could support patient stratification, identify individuals who are more likely to benefit from specific interventions, and guide the development of personalized treatment strategies. Ultimately, integrating multimodal biomedical data with interpretable AI may facilitate biomarker discovery and contribute to improved diagnosis, therapeutic development, and treatment monitoring for AD and other age-related neurological disorders.

Future work can focus on expanding the dataset to include larger and more diverse populations, incorporating longitudinal data to model long-term treatment effects, and exploring alternative feature selection strategies that further improve robustness and interpretability. Currently, patients are predominantly Caucasian, which reflects the demographic of Columbia, MO, and the results of a diversified genotypic pool could offer a deeper insight into the impacts of rapamycin treatment [5]. Other ambitions include an ablation study to further cross reference these results with other methodologies along with analyzing brain regions to see the difference in baseline and post treatment activity.

## Data Availability

All data produced in the present study are available upon reasonable request to the authors.

https://github.com/jianlin-cheng/TreatmentFormer

## Author Contributions

Conceptualization, Jianlin Cheng and Ai-Ling Lin; AI methodology, Jianlin Cheng and Chris Wang; patient data: Ai-Ling Lin, Carter Woods; software, Chris Wang, Carter Woods, Thong Nguyen and Jian Liu; computational experiment and data collection, Chris Wang, Thong Nguyen, Jian Liu, and Carter Woods; validation, Chris Wang, Thong Nguyen, Carter Woods, Jian Liu, Ai-Ling Lin and Jianlin Cheng; writing, Chris Wang, Jianlin Cheng, Ai-Ling Lin, Carter Woods, Thong Nguyen, Jian Liu; all authors have read and agreed to the published version of the manuscript.

## Funding

This work was supported by a Curators’ professorship and the Paul K. and Diane Shumaker Professorship to J.C. and the MU Radiology and NextGen start-up funding to A.L.

## Institutional Review Board Statement

Not applicable.

## Informed Consent Statement

Informed consent was obtained from all subjects involved in the study.

## Data Availability Statement

The code and document of TreatmentFormer is available at: https://github.com/jianlin-cheng/TreatmentFormer

## Conflicts of Interest

The authors declare no conflicts of interest.

## Disclaimer/Publisher’s Note

The statements, opinions and data contained in all publications are solely those of the individual author(s) and contributor(s) and not of MDPI and/or the editor(s). MDPI and/or the editor(s) disclaim responsibility for any injury to people or property resulting from any ideas, methods, instructions or products referred to in the content.

## Notes

### Competing Interest Statement

The authors have declared no competing interest.

### Clinical Trial

05386914

### Author Declarations

The study used (or will use) ONLY openly available human data that were originally located at: Rapamycin enhances neurovascular, peripheral metabolic, and immune function in cognitively normal, middle-aged APOE4 Carriers: genotype-dependent effects compared to non-carriers.

